# ‘Exploring Nurses’ Perspectives on Video Consultations and the Nurse–Patient Relationship’

**DOI:** 10.64898/2025.12.14.25342235

**Authors:** Sissel Groth, Anne Brødsgaard, Signe Stelling Risom, Jens Dahlgaard Hove, Stine Rosenstrøm

## Abstract

**Aim:** This study aims to deepen our understanding of how outpatient clinic nurses experience the use of video consultations. Further, the prerequisites that outpatient clinic nurses find important to increase the usage of video consultations in outpatient clinics.

**Materials and methods:** A hermeneutic qualitative exploratory approach was utilised to investigate these experiences and perceptions comprehensively. Data were collected through two qualitative focus group interviews with a purposive sample of 12 nurses from two Danish cardiology outpatient clinics.

**Results:** Data provided insights into the perspectives of nurses, resulting in three main themes: *Video consultations must be used at the right time*, *Nursing care must remain as a relational practice*, and *Missing initiatives for enabling nurses to use video consultations*. The study highlights the importance of organisational support and infrastructure in shaping nurses’ attitudes toward video consultations. Strategies for successful implementation include targeted training programmes, user-friendly technology, and a well-defined approach.

**Public Contributions:** No Patient or Public Contribution

## 1. Introduction

The evolution of digital technologies has introduced new terminologies and operational considerations (WHO, 2022). Virtual health care has rapidly become vital to how health care is delivered, received, and experienced (Pu et al., 2023). In its guidelines and recommendations, the WHO prioritises client-to-provider and provider-to-provider telemedicine as two of the most important and influential digital technologies for strengthening the healthcare system (WHO, 2019). Increasing demand for outpatient appointments is a global challenge in the healthcare system, and digital solutions such as video consultations may help meet those challenges (Morrison et al., 2021). Although the COVID-19 pandemic provided enormous impetus for healthcare services across all sectors to transition to virtual healthcare alternatives in a short amount of time (Pu et al., 2023), the maintenance seems challenging (Ganguli et al., 2023). Virtual health care, such as video consultations, are increasingly used as part of the digitisation of health care and can potentially improve the quality of care and increase patient accessibility (Golbus et al., 2023; Khanassov et al., 2024).

A systematic review revealed that nurses working with video consultations and other technology in caring for patients with cardiac disease experienced a sense of insecurity derived from different internal and external influences, which challenged the use of technology in health care (Rosenstrøm et al., 2024). Healthcare providers may percept technology usability as challenging, especially for older patients, which is a barrier for implementation of technology in health care, and may be reason for the lack of maintenance use of video consultations (Khanassov et al., 2024). A review focusing on contextual factors related to the implementation of video consultations found that patients generally respond positively, recognising the benefits and opportunities they provide (Aidemark, 2022). From the patient’s perspective, the most common advantages include saving time and avoiding the need to travel (Aidemark, 2022).

Telehealth is characterised by a complex interplay of technological, social, and organisational factors, where success of the technology depends on the ‘match’ between these elements (Sharma & Clarke, 2014). Since nurses play a central role in the healthcare system, and in each of the above factors, their experiences of different perspectives of video consultations can provide important insight into how this technology affects patient care, work routines, and professional interaction (Wasik, 2020). Further, how nurses experience and adapt to this new form of communication to offer video consultations as politically intended in the health care system (Borsch et al., 2024). Ideally, virtual healthcare initiatives such as video consultations should be carefully planned, and their barriers and facilitators should be assessed before they are implemented or scaled (Pu et al., 2023). This planning process supports delivering quality virtual health care and maintains the relevance of virtual care initiatives in an increasingly digital future (Pu et al., 2023).

Understanding the barriers that outpatient clinic nurses experience when offering and using video consultations and their perspectives of how it affects nursing care is important knowledge for the ability to eliminate the barriers, accommodate the prioritises in clinical practice, and leverage the opportunities video consultations offer all adult patients in the outpatient clinic.

## 2. The study

### 2.1. Aim and objective

This study aims to deepen our understanding of how outpatient clinic nurses experience the use of video consultations. Further, the prerequisites that outpatient clinic nurses find important to increase the usage of video consultations in outpatient clinics.

## 3. Material and methods

### 3.1. Design and theoretical framework

Given the study’s objectives to shed light on outpatient clinic nurses’ experiences and perceptions, a qualitative exploratory research method was used (Polit & Beck, 2017). The study adopts a hermeneutic approach, in which outpatient clinic nurses’ lived experiences are used to interpret and understand individual experiences (Polit & Beck, 2017). The hermeneutic approach recognises that the researchers’ pre-existing knowledge and understanding of the studied phenomena influences their interpretation of the findings, which is achieved by continuously moving between the parts and the whole of the data being analysed (Polit & Beck, 2017). The reliability of this study was ensured through descriptions using Lincoln and Guba’s criteria for trustworthiness during the analysis process (Krefting, 1991). The Consolidated Criteria for Reporting Qualitative Research (COREQ) checklist was followed throughout the study’s development, execution, and reporting phases (Tong et al., 2007).

### 3.2. Data collection

#### 3.2.1 Sampling and recruitment

The study was carried out in a university hospital located in the capital region of Denmark, specifically within a cardiology outpatient clinic spread across two sites in Copenhagen. Two focus group interviews were conducted, one in each location familiar to the participants. The focus group interviews were conducted to gather information from the different participants’ point of view (Stalmeijer et al., 2014). Further, to understand and explain the meanings, beliefs, and cultures that influence the participants feelings, attitudes, and behaviours (Stalmeijer et al., 2014). The groups were designed to include five to eight participants: small enough to allow each individual to participate fully and be heard, yet large enough to allow for varying opinions and perspectives (Stalmeijer et al., 2014).

The participants in each focus group were all female nurses working in cardiological clinical practice and knew each other from their work (Table 2). All were knowledgeable about video consultations, having been introduced to the technology and trained on its use through a partner training session during the implementation process in 2021. This training included a brief technical introduction, instructions on scheduling video consultations, and operational guidance. A flowchart was created to help clinicians determine the suitability of video consultations. Although video consultations were prioritised for implementation at the time of the interviews, their use was optional in both outpatient clinics, and in-depth experience was not required.

All outpatient clinic nurses were equipped with headphones and a video camera connected to their computers in the consultation rooms. For video consultations, nurses connected with patients via an e-health platform, which required a link accessible through a smartphone, tablet, or computer. Other than connecting via a video link on the health platform, video consultations necessitated the same preparatory work as physical consultations. While video consultations were accessible to all participants, they had not been implemented to the same extent as physical or telephone consultations.

The participants’ experience with the use of video consultations was not a requirement for participation in the focus group interview, but instead the interest in sharing their thoughts on the use of video consultations.

#### 3.2.2. Data sources/collection

The first and last author, who was responsible for conducting the interviews, participated in a pre-understanding interview (Tufford & Newman, 2012) conducted by the third author. This preunderstanding interview aimed to reveal the author’s preunderstanding of the topic, thereby promoting explicitness and transparency during the data collection and processing stages (Tufford & Newman, 2012). Both authors possess an in-depth experience as clinical registered nurses in the cardiological field. The first author has no clinical experience with outpatient clinic treatment or virtual consultations. The last author has long-standing experience with outpatient clinic treatment and virtual consultations. The first author moderated both interviews and thus had the responsibility to facilitate discussions and exchange of ideas between the participants (Stalmeijer et al., 2014). The last author ensured that all themes were covered through the questions from the interview guide (Table 1) and that all participants could express their views (Krueger, 2006).

**Table 1.** Interview guide.

| Themes | Questions |
| --- | --- |
| **Attitude towards IT solutions** | What are your perceptions of physical consultations being replaced by virtual ones?<br>Can you unfold pros and cons when using digital solutions such as video consultations in the outpatient clinic? |
| **Functional and clinical needs** | When is a consultation successful to you?<br>What does a successful consultation mean to you? |
| **Emotional needs** | How do you feel during a consultation?<br>What do you value?<br>How do you feel about physical consultations being replaced with virtual ones? |
| **Social needs** | How would you describe your relationship with patients during a consultation? |

**Table 2.** Participant characteristics.

| **Interview** | **Current position** | **Clinical experience from cardiac outpatient clinic practice** | **Cardiological field** | **Number of video consultations (estimated by the participants)** |
| --- | --- | --- | --- | --- |
| **1**<br>(Participant 1-7) | Registered nurses (n=7)<br>Advanced practice nurses (n=0)<br>Staff nurses (n=0) | Mean 9 years<br>Range 1-15 years<br>Median 10 years | Heart rehabilitation, heart valve patients, treatment for hypertension, arrhythmia, heart failure, endocarditis, anticoagulation treatment, and postpartum patients | From 1 to 50 (one participant had conducted approximately 200, as she had completed the virtual postpartum programmes in the outpatient clinic) |
| **2**<br>(Participant 8-12) | Registered nurses (n=4)<br>Advanced practice nurses (n=0)<br>Staff nurses (also RN) (n=1) | Mean: 12 years<br>Range: 4-22 years<br>Median 8 years | Heart rehabilitation, heart valve patients, treatment for hypertension, arrhythmia, heart failure, and anticoagulation treatment | From 0 to 20 |

The interview guide was created based on a systematic literature review, clinical experiences, and qualifications of the first and last authors. The interview guide served as an inspiration to unpack the participants’ experiences and perceptions. It was created to uncover what was deemed valuable in patient-provider relationships, explore different perspectives and requirements of physical consultations, identify perceived limitations, and reflect upon the potential of video consultations. It was emphasised that reaching a consensus was not the goal; rather, the aim was to encourage the participants to share their experiences, perspectives, and curiosities with one another (Krueger, 2006). All themes and related questions were thoroughly discussed. While the questions did not always follow the interview guide’s chronological order, the guide was instrumental in ensuring that all themes were illuminated to sufficient depth and nuance. The interviews were initiated by framing and clarifying the study’s aim and progressively focusing the participants’ attention on the topics of most significant interest (Krueger, 2006). The facilitator of the interviews tried to take advantage of the group homogeneity in terms of educational background and current context in facilitating open communication, exchange of ideas, and a sense of safety in expressing conflicts of concerns (Stalmeijer et al., 2014).

Each session lasted approximately 90 minutes and was audio-recorded. The interviews were transcribed verbatim by the first author. The names of the participants were anonymised and replaced with numbers for analysis and reporting.

### 3.3. Ethical considerations

The interview dates were chosen by the first and last author, and all the nurses in the outpatient clinic were invited to participate in the focus group interviews, totalling 17 clinical nurses. Four invited nurses cancelled due to having scheduled time off, and one due to being too busy on the interview day. Participation in the study was voluntary and took place during working hours. Both written and oral information about the study was provided to the participants, and written informed consent was obtained from all participants before starting the interviews. The study did not require approval from Danish research ethics committees (Journal No. F-23075094).

### 3.4. Analysis

The transcribed interviews were analysed using thematic analysis, for systematically identifying, organising, and gaining insights into patterns of meaning or themes across the interview data (Braun & Clarke, 2006). The thematic analysis acknowledges the active role of the researcher in identifying patterns/themes, deciding on their relevance, and reporting them to the reader (Braun & Clarke, 2006). The process begins as researchers identify potential patterns of meaning and issues of interest that can occur during data collection. The analysis involves constantly moving back and forth between the entire dataset, the coded extracts of data being analysed, and the ongoing analysis of the data being produced (Braun & Clarke, 2006). The hermeneutic approach was used to facilitate a deeper understanding of the interview texts by alternating between parts of the data and the dataset as a whole, interpreting the findings by continually questioning their meaning (Polit & Beck, 2017). The analysis followed six phases, as recommended by Braun and Clarke (2006):

Phase 1: *Familiarising oneself with the data* implied transcribing the data as an interpretative act, reading and re-reading the data, and searching for meanings and patterns.

Phase 2: *Generating initial codes* entailed a systematic examination of the entire dataset, paying full and equal attention to each item and identifying aspects that might form the basis of repeated patterns. The coded data were organised into meaningful groups relative to the study’s aim.

Phase 3: *Searching for themes* involved sorting the initial codes into potential themes, with all relevant coded data extracts collated within these identified themes. A mind map was used to create an overview and understand the connections between themes, using separate pieces of paper with the name of each code and brief descriptions to experiment and organise themes. Codes that did not seem to belong anywhere were collected in a ‘miscellaneous group’.

Phase 4: *Reviewing themes* required refining the themes through critical reviewing. First, we considered whether the codes formed a coherent pattern and adequately captured the perspectives within the coded data. Second, reading alternating sections of the text generated new understandings of the individual sections and the text as a whole. The refinement process continued until no substantial new insights were added.

Phase 5: *Defining and naming themes* involved identifying the ‘essence’ of each theme and determining what aspect of the data each theme captured.

Phase 6*: Producing the report* entailed reporting the reasons for theoretical, methodological, and analytical choices, describing the coding and analysis processes in sufficient detail to capture the essence of the points without unnecessary complexity, and providing a thorough description of the context to ensure transferability. Quotations from the interview text were used to illustrate the analytical process, supporting the transparency and trustworthiness of the findings and the authors’ interpretations of the data (Table 3 in the appendix). Researcher triangulation was conducted by the first and last author to validate the analysis throughout the process. In this phase, the entire research team completed peer debriefing and researcher triangulation to validate the interpretation.

**Table 3.**
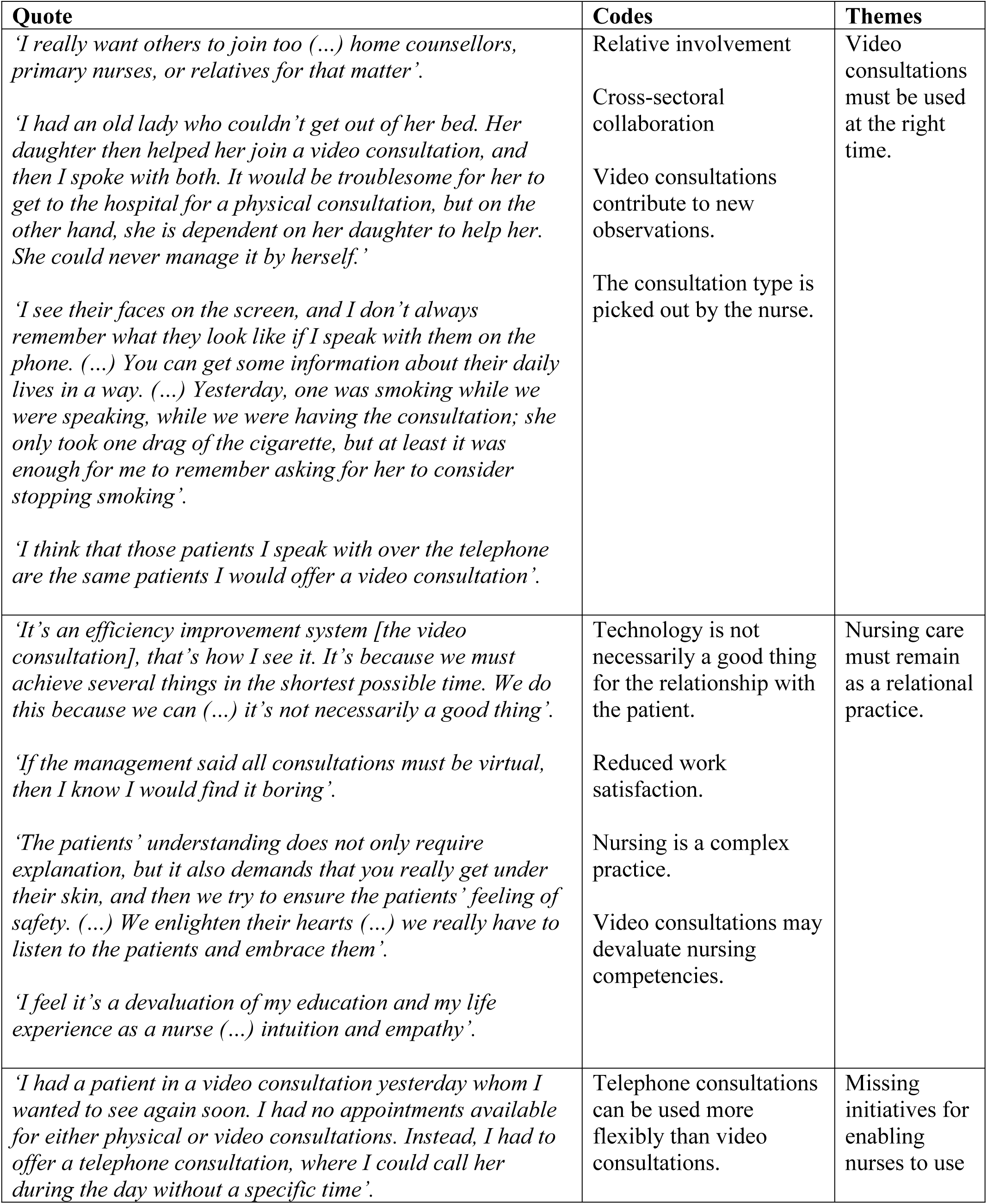
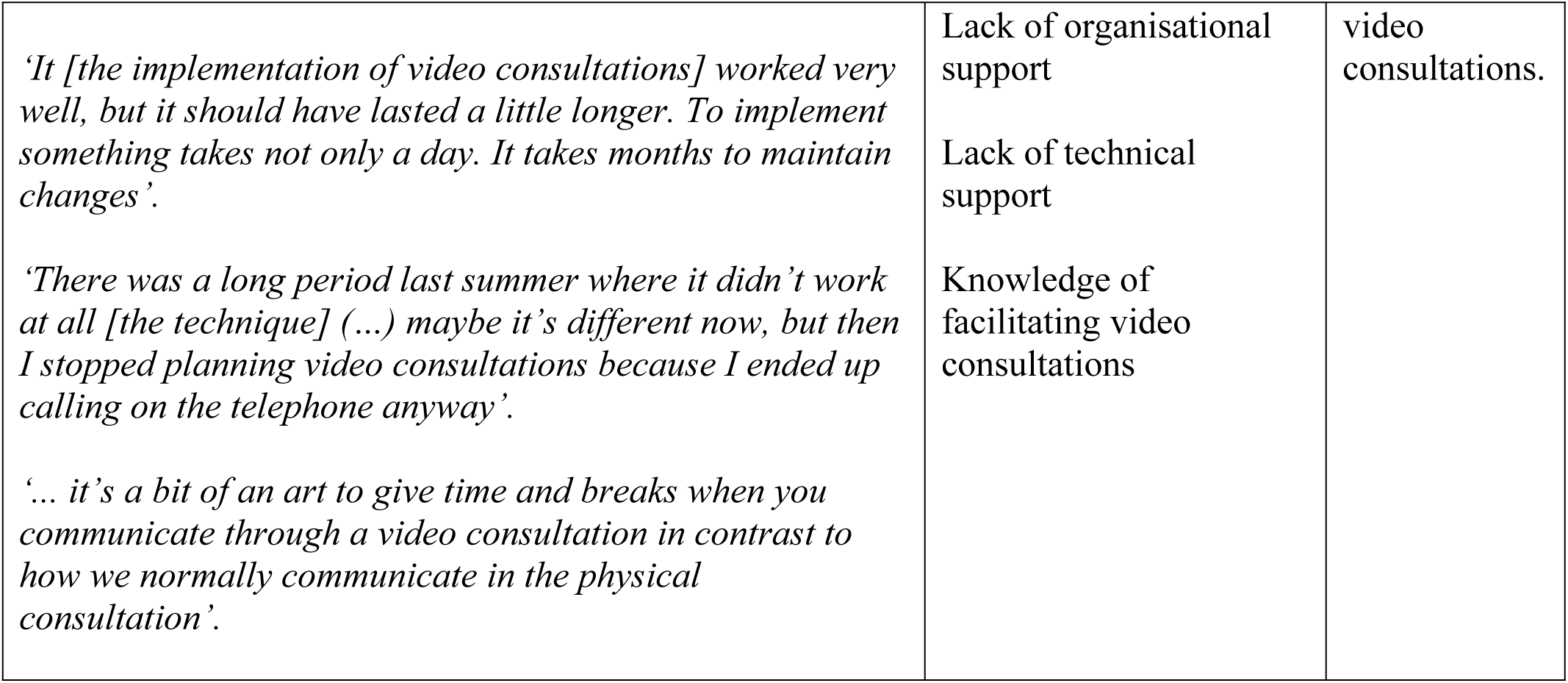
Example of coding.

## 4. Results

The analysis of data resulted in three themes based on 12 codes: 1) *Video consultations must be used at the right time*, 2) *Nursing care must remain as a relational practice*, and 3) *Missing initiatives for enabling nurses to use video consultations*. These themes are presented in Table 3, supported by illustrative, representative, and rich quotations.

### 4.1. Video consultations must be offered at the right time

A common view was an overall concern about the increasing use of technological solutions in outpatient clinic treatment and care activities. In general, the participants saw both possibilities and limitations in the different types of consultations: physical, telephone, and video. When choosing among these types, both patients’ and nurses’ preferences were considered important factors. One participant noted:

> *‘I think that those patients I speak with over the telephone are the same patients I would offer a video consultation’. (Participant 12)*

The participants acknowledged the nurses’ role in initiating the use of technology and how the different consultation types are presented to patients. All agreed that video consultations should be carried out if patients demand or prefer them:

> *‘Whether it is the choice between attendance [physical consultation] or video consultation, I think they should be allowed to choose (…) I think it would be a shame if they were forced to do video just because we believe it is smart’. (Participant 9)*

The participants expressed that the first meeting with a patient should preferably be a physical consultation because it establishes the relationship. They acknowledged that subsequent nursing consultations could be conducted effectively via video, offering value to some patients. The participants also noted that video consultations could be an efficient means of collecting additional information about the patient’s daily life, thus serving as a good alternative to telephone consultations by supporting their memory of the patient:

> *‘I see their faces on the screen, and I don’t always remember what they look like if I speak with them on the phone. So, I see their faces, and I enter their living room for better or worse. (…) You can get some information about their daily lives in a way. (…) Yesterday, one was smoking while we were having the consultation; she only took one drag of the cigarette, but at least it was enough for me to remember asking her to consider stopping smoking’. (Participant 2)*

The participants highlighted that the choice of consultation type (physical, telephone, or video) should be tailored to the specific needs of each patient:

> *‘I had an old lady who couldn’t get out of her bed. Her daughter then helped her join a video consultation, and then I spoke with both. It would be troublesome for her to get to the hospital for a physical consultation, but on the other hand, she is dependent on her daughter to help her. She could never manage it by herself’. (Participant 2)*

However, others emphasised that the use of video consultations could initiate a stronger cross-sectoral collaboration by including home care nurses and general practitioners in these consultations to collectively support vulnerable patients. Video consultations could also be used to involve relatives who might not attend physical consultations. There was a consensus that involving the primary care sector and relatives could be highly beneficial for patients but would require further development and integration into care and treatment workflows to monitor specific care and treatment tasks and system access.

### 4.2. Nursing care must remain as a relational practice

The participants expressed that they found it difficult to justify the use of video consultations over physical ones:

> *‘It’s an efficiency improvement system [the video consultation], that’s how I see it. It’s because we must achieve several things in the shortest possible time. We do this because we can (…) it’s not necessarily a good thing’. (Participant 1)*

The perceived nurse shortage across the healthcare system influenced the nurses’ perceptions and perspectives on implementing video consultations. Nurses are aware that resources must be distributed and utilised wisely. The participants felt that the political agenda and arguments supporting telemedicine, and the benefits of video consultations had not been clearly communicated by the department or hospital management, leading to reduced motivation to adopt video consultations. Furthermore, the participants agreed that video consultations do not save time but rather degrade the quality of nursing care by challenging traditional patient interactions. The participants feared that video consultations would diminish their job satisfaction:

> *‘If the management said all consultations must be virtual, then I know I would find it boring’. (Participant 8)*

As the quote illustrates, the participants felt video consultations could hinder establishing meaningful patient relationships. Nonetheless, they were aware of the necessity of keeping pace with technological advancements in society to meet patients’ evolving demands and expectations. The participants also expressed the need to acquire new knowledge and skills to adapt effectively. The participants were strongly committed to providing nursing care that ensures and maintains patient empowerment, safety, and security. More specifically, all participants felt a great responsibility to support and encourage their patients to manage their heart disease effectively. One participant expressed:

> *‘We must educate the patients to react to symptoms, take care of themselves, and prevent (…) the aggravation of heart failure. We must guide them in weight loss, diet, physical activities, and lifestyle risk factors, explaining the importance of taking the prescribed medicine’. (Participant 4)*

A common view was that the desired achievements in nursing consultations required physical contact to establish a meaningful relationship with the patients. From the participants’ perspective, a meaningful relationship required ‘good chemistry’, empathy, and intuition:

> ‘*The patients’ understanding does not only require explanation, but it also demands that you really get under their skin, and then we try to ensure the patients’ feeling of safety. (…) We enlighten their hearts (…) we really have to listen to the patients and embrace them’. (Participant 5)*

A participant described feeling that video consultations diminished the value of her education and clinical experience by undercutting her professional competencies:

> *‘I feel it’s a devaluation of my education and my life experience as a nurse (…) intuition and empathy’. (Participant 1)*

The participants felt a strong sense of responsibility towards their patients and thus had to advocate for the most suitable consultation type.

### 4.3. Missing initiatives for enabling nurses to use video consultations

The participants emphasised that management had an important role in directing how nurses and other healthcare professionals should prioritise video consultations. Some participants were concerned that video consultations would only increase the nurses’ workload with additional tasks, such as teaching patients to use the technology, dealing with technical issues, and meeting a demand for greater availability and flexibility. When patients needed earlier appointments than scheduled, participants found it more straightforward to offer a telephone consultation rather than a video consultation because it was easier to integrate into an already busy schedule:

> *‘I had a patient in a video consultation yesterday whom I wanted to see again soon. However, I had no appointments available for either physical or video consultations. Instead, I had to offer a telephone consultation, where I could call her during the day without a specific time’. (Participant 2)*

The organisation of the outpatient clinics and scheduling of patients sometimes acted as barriers to video consultations because only telephone consultations could be flexibly added as an extra option. The participants stated that management wished for video consultations to be offered equally alongside physical and telephone consultations in the outpatient clinic. Furthermore, management needed to prioritise time for education and the development of digital competencies to ensure high-quality video consultations:

> *‘It is difficult to make changes when there isn’t time for it. (…) What is it, really? What should I do when I have a video [consultation]? It’s very rare that we have enough time, and then you don’t take ownership of it because it [the implementation process] feels a little sloppy, and then you become sloppy, too’. (Participant 12)*

At the beginning of the video consultation implementation process, the participants received partner training to become familiar with the technology. However, they mentioned that it was too brief:

> *‘It [the implementation of video consultations] worked very well, but it should have lasted a little longer. To implement something takes not only a day. It takes months to maintain changes. (Participant 12)*

After the partner training, the participants felt they lacked support in addressing questions and challenges related to video consultations. They argued that adequate resources must be allocated to training staff to maintain motivation, given that outpatient clinic nurses’ time was scarce and utilised to the maximum:

> *‘There was a long period last summer where it didn’t work at all (…) maybe it’s different now, but then I stopped planning video consultations because I ended up calling on the telephone anyway. (…) If you have to discover yourself, that it doesn’t work [the video link] when you are sitting with the patient, then the motivation disappears’. (Participant 11)*

The participants explained that they often used telephone consultations when video links did not work, leading many to prefer telephone consultations. It was a commonly held belief that video consultations require different communicative competencies to maintain good relationships with patients compared to telephone or physical consultations. The participants explained how video consultations demand alternative approaches to understanding patients’ perspectives and symptoms when not all senses can be engaged in the overall assessment of the patient. Additionally, facilitating video consultations requires different approaches compared to physical consultations:

> *‘… it’s a bit of an art to give time and breaks when you communicate through a video consultation in contrast to how we normally communicate in the physical consultation’. (Participant 2)*

Beyond the need for education on conducting video consultations for nurses, the participants also highlighted the necessity of educating patients on how to use video consultations effectively. In current practice, patients’ technical support needs fall on the nurses, with time allocated for this support detracting from the consultation time, thus necessitating a prioritisation of technical support over health-related discussions. Looking ahead, the participants anticipated that telemedicine would face far fewer challenges and barriers:

> *‘I think it will change automatically. Seniors, including myself, have a little fear of it [video consultations] because we didn’t grow up with it. (…) I’m afraid that the technology won’t work’. (Participant 7)*

Despite anticipating that the demand from future patients may lead to the natural adoption of video consultations, the participants agreed on the importance of being proactive in preparing themselves for these future demands.

## 5. Discussion

The findings provided an in-depth understanding of the factors that can affect video consultations, which, when unaddressed, contribute to the limited scope of video consultations in outpatient clinic settings. The finding that ‘*Video consultations must be used at the right time*’ underscored nurses’ recognition of the challenges and limitations, as well as the potential benefits of video consultations for both nurses and patients. In this study, nurses appreciated the convenience of video consultations offered to patients and their families by saving transportation time and allowing more flexible scheduling. Video consultations convenience aligns with findings from another study, which reported video consultations as significant time and cost savers for patients (Donaghy et al., 2019). The participants also viewed video consultations as an opportunity to enhance cross-sectoral collaboration with home care nurses and general practitioners, supporting the valuable exchange of useful knowledge for improved patient care. Similarly, other studies highlighted the benefits of cross-sectoral video consultations in contributing to patient-centred care, the clarity of roles, treatment continuity, and patient satisfaction (Jensen et al., 2024; Trabjerg et al., 2021). Some nurses noted that video consultations offered them opportunities to gather additional information about their patients by seeing them in their home environments, in contrast to telephone consultations. Additionally, video consultations enable nurses to pick up on non-verbal cues during patient assessments, which is not feasible with telephone use alone (Donaghy et al., 2019). The nurses acknowledged their role in initiating video consultations based on what they deemed best for their patients, potentially positioning patients in a passive role (Molina-Mula & Gallo-Estrada, 2020). Furthermore, the nurses acknowledged that patients’ needs, and demands are highly important when choosing the type of consultation. The nurses’ clinical experience working in a cardiology outpatient clinic setting varied, which could impact their use of video consultations. In addition, one study found no correlation between age or seniority and the intention to use telenursing and video consultations (Kats & Shmueli, 2024).

The insight that *Nursing care must remain as a relational practice* revealed the importance that the nurses placed on their relationship with patients to deliver high-quality care. The nature of the clinician-patient relationship directly affects the quality of care (Molina-Mula & Gallo-Estrada, 2020). It has been noted that a good relationship initiated by the expert professional directs knowledge, professional experience, and clinical skills toward the specific needs of each patient (Molina-Mula & Gallo-Estrada, 2020). These findings align with the nurses’ perceptions in this study, emphasising the value of physical interaction. Professional judgment, seen as an interplay of senses, involves being present with one’s entire personality and professionalism (Martinsen, 2017).

The nurses found achieving this presence challenging without physical interaction. Referencing Løgstrup (1956/1986), care ethics suggest that humans are inherently connected and have a moral obligation to act in others’ best interests and care for the lives entrusted to them (Delmar, 2012). The nurses in this study expressed concerns about fulfilling their ethical and moral responsibilities through video consultations. It has been shown that practising care by telephone is complex due to the inability to offer physical reassurance, such as placing a comforting hand on the patient (Boström et al., 2020; Ladin et al., 2021). The nurses in this study expressed the feeling that video consultations could devalue their education and clinical experience by challenging their intuition and empathy. The potential devaluation of nurses’ clinical expertise through the adoption of video consultations was also highlighted as a significant concern in adapting to this technology (Sharma & Clarke, 2014).

The finding of ‘*Missing initiatives for enabling nurses to use video consultations’* revealed that managerial prioritisation and direction were crucial in facilitating the use of video consultations. Managerial encouragement, adequate training, and the provision of resources were found to be imperative for enabling nurses to employ video consultations in remote patient care (Kats & Shmueli, 2024). One study identified the willingness of institutional forces at both societal and organisational levels to invest in strategic purpose and funding as the driving force behind successful implementation (Aidemark, 2022). The participants in this study were all introduced to video consultations through the same implementation process. Nevertheless, they incorporated video consultations into their clinical practice to varying extents. Rogers (2003) noted that individuals within a social system do not adopt new ideas simultaneously, necessitating diverse communication strategies during implementation. The participants pointed out the need to develop their technological skills before offering video consultations. The focus on enhancing competencies and relatedness is supported by several studies (Boström et al., 2020; Deci & Ryan, 2008).

Reflecting on theory and their practice allows nurses to redefine their professional roles to provide remote patient care while ensuring the delivery of high-quality care (Boström et al., 2020). Moreover, enhancing competencies is essential for fostering a feeling of autonomy, which is vital for intrinsic motivation in humans (Deci & Ryan, 2008). Successful implementation relies on motivated individuals to come up with innovative solutions to solve problems as they arise (Lu et al., 2021). Therefore, the sustainable implementation of video consultations requires iterative processes, involving stakeholders to ensure engagement, process mapping to delineate the inputs, outputs, and steps of a specific change initiative, and problem-solving (Lu et al., 2021).

### 6.1 Strength and limitations of the work

This study has a number of strengths and limitations. The fact that the interviewers was part of the same department as the participants presented a limitation in achieving objectivity in conducting the interviews. Uncovering the interviewers preconception through preunderstanding interview sharpened the awareness of tacit knowledge. At the same time, familiarity with the department, the culture, and the organisation enabled the interviewer to probe further and unravel the nuances in statements. To ensure credibility, the choice of facilitator was based on having the least familiarity with the outpatient clinic and its nursing staff. A significant strength was the number of participations, which were representing over half of the outpatient clinic nursing staff (totalling 17). The interview process and techniques were consistent across the interviews, focusing on exploring different subjects and perspectives until data saturation was achieved. To ensure the study’s transferability, detailed descriptions of the data were provided. Rich descriptions of the qualitative methodological approach strengthened the study’s dependability, enabling the exploration of the research aim and showing that the findings were consistent. Coding accuracy and reliability were measured by the research team. A high level of reflexivity throughout the analytical process through continuous discussions among the authors strengthened the study’s confirmability. Furthermore, conducting a preunderstanding interview before the two focus group interviews made the researchers more aware of their roles and preconceptions. Finally, the study’s confirmability was strengthened through a rigorous and systematic presentation of the study processes, which was confirmed continuously through researcher triangulation.

### 6.2. Recommendations for further research

The findings of this study indicate unmet needs and suggest areas that may lead to better implementation of video consultations in outpatient clinic settings. Based on this knowledge, recommendations for guidelines can be proposed. To ensure successful and sustainable implementation, management must foster a motivated workforce capable of developing innovative solutions to emerging challenges. Management must ensure that staff members possess the knowledge, skills, and attitudes to support the integration of video consultations on par with telephone and physical consultations. Thorough and systematic training of nurses is necessary, along with a sustained focus on identifying and addressing the opportunities and challenges presented by the technology in relation to patient care.

## 7. Conclusion

In conclusion, the nurses in this study found video consultations advantageous for some patients, their families, and the healthcare system. The present work contributes to a deeper understanding of the factors influencing outpatient clinic nurses’ perceptions and motivation toward using video consultations. The themes *Video consultations must be used at the right time*, *Nursing care must remain as a relational practice*, and *Missing initiatives for enabling nurses to use video consultations* reveal that the nurses perceive video consultations as potentially diminishing the quality of the care they provide, which they are unwilling to compromise. Nursing care is rooted in nurses’ moral and ethical responsibility towards their patients. While acknowledging the potential benefits of video consultations for some patients, the nurses emphasised that the choice of consultation type must be tailored to individual needs. Addressing these needs requires managerial and organisational support and prioritisation in terms of resources and education for both clinicians and patients.

## Data Availability

All relevant data are within the manuscript and its Supporting Information files.

## 8. Acknowledgements

We are grateful to all the nurses from the outpatient clinics who participated in the study and the management of the cardiology department for their support of this study. Furthermore, we extend our gratitude for the funding.

